# Impact of a machine learning-powered algorithm on pathologist HER2 IHC scoring in breast cancer

**DOI:** 10.1101/2025.02.04.25321335

**Authors:** John Shamshoian, Zahil Shanis, Ryan Cabeen, Limin Yu, Shreya Chakraborty, Marc Thibault, Blake Martin, Harshith Pagidela, Dinkar Juyal, Syed Ashar Javed, William Qian, Juhyun Kim, Ylaine Gerardin, Beckett Rucker, Jacqueline Brosnan-Cashman, Harsha Pokkalla, Jimish Mehta, Amaro Taylor-Weiner, Eric Walk, Andrew Beck, Michael C. Montalto, Ben Glass, Santhosh Balasubramanian

**Affiliations:** PathAI, Boston, MA

## Abstract

**Background:** HER2 expression level is a key factor in determining the optimal treatment course for breast cancer patients. Roughly 15% of breast cancers are HER2(+), and determination of HER2 status is routinely assessed by immunohistochemistry (IHC). Accurate assessment of the HER2 IHC score by pathologists is therefore critical, especially in light of novel therapeutic approaches demonstrating efficacy in the HER2-low setting. However, there is an opportunity to improve inter-pathologist agreement at the lower levels of HER2 scoring (0, 1+, and 2+).

**Methods:** A machine learning model (AIM-HER2) was developed to generate accurate, slide-level HER2 scores aligned with ASCO-CAP guidelines in clinical breast cancer HER2 IHC specimens. AIM-HER2 was assessed as an AI-assist tool in a retrospective reader study, where 20 HER2-trained pathologists scored breast cancer cases (N=200) with and without AIM-HER2 assistance using a 2-cohort crossover design with a 3-week washout. A separate panel of 5 expert HER2 pathologists read all 200 cases manually to establish reference scores.

**Results:** In a significant fraction of cases examined, less than 70% inter-pathologist agreement was observed. When used as an AI assist tool, AIM-HER2 improved inter-rater agreement overall and specifically at the 0/1+ and 1+/2+ cutoffs. Similarly, AIM-HER2 AI-assist significantly increased PPA at the 0/1+ and 1+/2+ cutoffs. When interacting with the AI-assist tool, pathologists displayed a wide range of override rates, and the quality of a pathologist’s overrides was correlated with their manual accuracy. Lastly, the impact of the reference panel on AIM-HER2 accuracy metrics was assessed, revealing that measurements of model accuracy are highly dependent on reference panel composition.

**Conclusions:** The use of AIM-HER2 as an AI-assist tool for scoring HER2 IHC in breast cancer may improve pathologist reproducibility and accuracy, particularly at the 0/1+ and 1+/2+ cutoffs.

## Introduction

The human epidermal growth factor receptor 2 (HER2), a tyrosine kinase receptor with pleiotropic downstream effects, is amplified in roughly 15-20% of breast cancers; elevated HER2 expression contributes to tumor progression and poor prognosis^1–3^. That said, therapeutic targeting of HER2 has long been a successful treatment strategy for patients whose tumors express elevated levels of HER2, with trastuzumab showing efficacy in early precision medicine studies^4^.

Due to the ability of agents such as trastuzumab to target HER2, testing of breast cancer specimens for HER2 expression is routine using HER2 immunohistochemistry (IHC) and, in some cases, fluorescent in situ hybridization (FISH). The American Society of Clinical Oncology (ASCO)/College of American Pathologists (CAP) have put forth guidelines for scoring HER2 immunohistochemistry based on the completeness and intensity of HER2 staining^5–7^. Under these guidelines, cases with either no HER2 expression or with partial, faint expression in fewer than 10% of cells are scored as HER2 0, cases with partial, faint expression of HER2 in greater than 10% of cells are scored as HER2 1+, cases with complete, weak-moderate HER2 expression in greater than 10% of cells are scored as HER2 2+, and cases with complete, intense HER2 expression in greater than 10% of cells are scored as HER2 3+. Cases scored as HER2 2+ are seen as equivocal and are reflexed to FISH testing to further assess the presence of *HER2* gene amplifications. Previous interpretations of HER2 scoring binarized patients into HER2-positive (HER2 3+ and HER2 2+/FISH+) and negative (HER2 0, 1+, 2+/FISH-) for eligibility for trastuzumab^8^. However, recent evidence from the DESTINY-Breast 04 trial has suggested that patients with HER2 1+ and HER2 2+/FISH-may have prolonged progression-free survival when receiving trastuzumab deruxtecan compared to chemotherapy ^9^.

Despite the long-standing guidelines for scoring HER2 positivity in breast cancer specimens and good concordance between pathologists for HER2-positive cases, pathologists show large degrees of intra- and inter-reader variability in scoring HER2, particularly at the 1+/2+ cutoff and in cases with a high degree of HER2 expression heterogeneity^10–14^. As such, it is estimated that 4% of negative cases and 18% of positive cases receive an incorrect score^15, 16^. Especially in light of the movement away from a dichotomized HER2 positive and negative classification system to include HER2-low as a distinct classification, approaches to improve the accuracy of HER2 scoring are necessary to maximize the benefit of precision therapeutics.

Artificial intelligence (AI) approaches have shown potential for aiding the pathology workflow, including for automated scoring of IHC images to improve accuracy and reproducibility^17–23^. Importantly, AI-based assistance strategies, where pathologists use an AI algorithm to guide assessments , have been shown to improve pathologist performance for multiple tasks^24^. Therefore, AI-assist pathology tools have a unique advantage over the use of standalone AI models.

Here, we describe the development of a machine-learning model (AI Measure of HER2, “AIM-HER2”) to generate accurate, slide-level HER2 scores aligned with ASCO-CAP guidelines in breast cancer HER2 IHC specimens. We assessed the performance of AIM-HER2 as a stand-alone tool, as well as in a larger study to examine pathologist accuracy and inter-reader variability with and without use of AIM-HER2 as an AI-assist tool.

## Methods

### Ethics

This study was conducted in accordance with the Declaration of Helsinki and Good Clinical Practice guidelines. Anonymized breast cancer HER2 IHC specimens and corresponding digitized WSIs were obtained from Cleveland Clinic, Dana Farber Cancer Institute, and Johns Hopkins Hospital as part of a data sharing agreement between PathAI and each institution. As part of each respective agreement, the Cleveland Clinic Institutional Review Board, the Dana Farber Cancer Center Institutional Review Board, and the Johns Hopkins Medicine Institutional Review Board approved the use of these samples for this study.

### Model Development

AIM-HER2 Breast is for research use only and is not for use in diagnostic procedures. AIM-HER2 makes use of three sub-algorithms: an artifact detection model, a tissue segmentation and classification model, and a HER2 scoring model (**Figure 1A**). Image artifacts and in situ carcinomas were identified using previously trained artifact and tissue segmentation models and were excluded, leaving only regions of invasive carcinoma to be analyzed. The artifact model was trained on annotations (N=2,951,972) of whole slide images (WSIs; N=13,670) provided by pathologists or pathologist-trained contributors (N=157). The tissue model was trained on annotations (N=157,697) provided by board-certified pathologists (N=68) on a cohort of breast cancer slides stained with HER2 IHC (N=5044) obtained from three academic medical centers.

**Figure 1.**
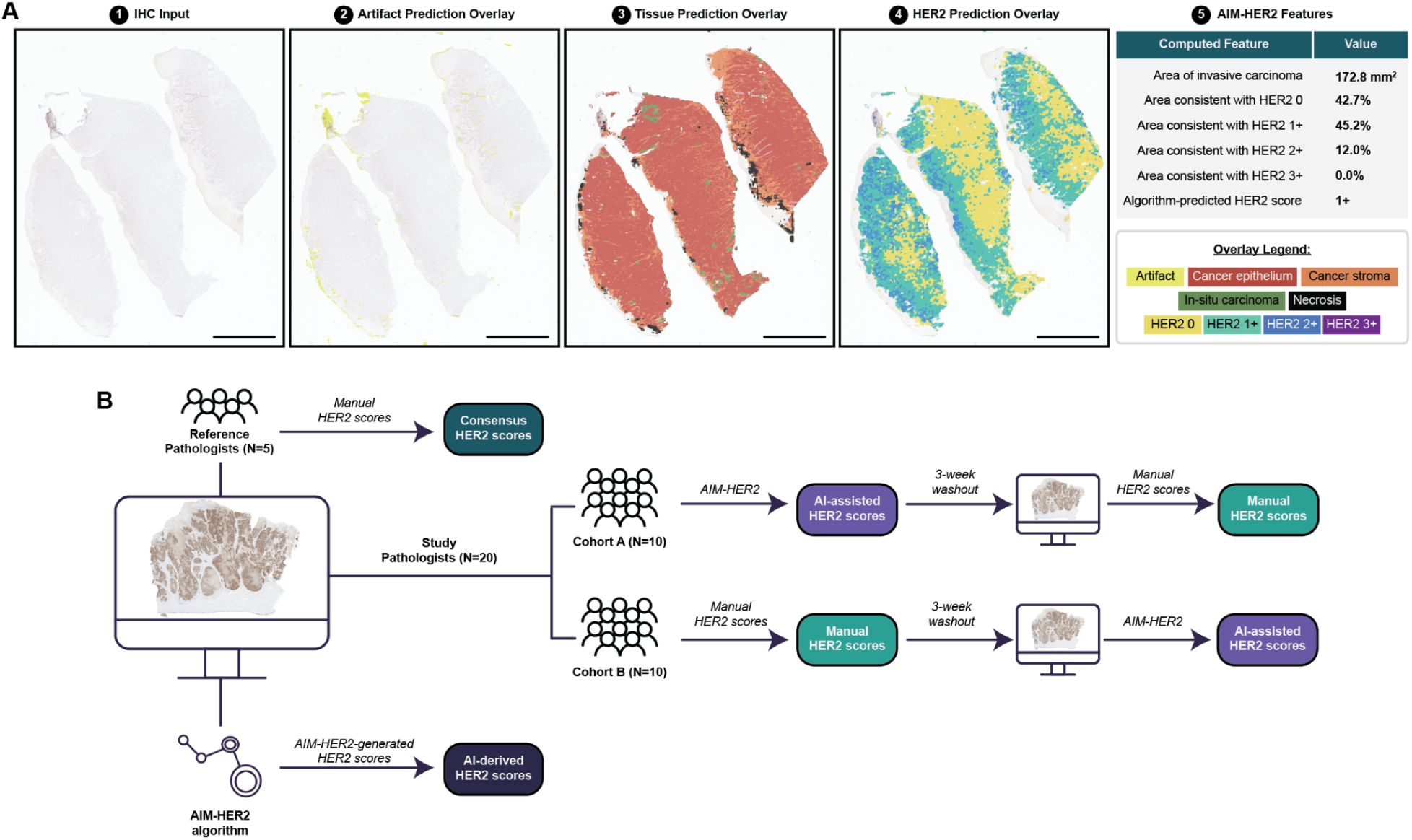
Study design. A) From WSI of HER2 IHC (panel 1), AIM-HER2 sub-algorithms detect and remove all artifact (panel 2), identify and classify regions of stroma, necrosis, and invasive & in situ cancer (panel 3), and identify cancer regions consistent with ASCO/CAP HER2 scores (panel 4). HER2 features are automatically generated, including a slide level HER2 score consistent with ASCO/CAP guidelines (panel 5). B) Study design for AIM-HER assisted read study. Pathologists were divided into two cohorts, who each scored both manually and with AI assistance using a cross-over design with a three week washout.

The HER2 scoring model utilized an additive multiple instance learning (aMIL) approach^25^ to predict HER2 scores directly from IHC WSIs and create interpretable heatmaps that depict HER2 predictions in tissue images (**Supplementary Figure 1**; **Figure 1A**). This model was trained using slide-level HER2 scores, leveraging regions predicted by the tissue model. The full slide set was divided into training (N=2,694), optimization (N=855), test (N=712), and standalone analytical verification and integrated analytical verification (N=783) sets. As part of this process, approximately 12,000 slide-level HER2 scores were obtained.

After model deployment on WSI of HER2 IHC, a model-generated overlay depicts the regions of tissue that match the ASCO/CAP definitions of each scoring level. Model outputs also include area of invasive carcinoma, areas consistent with each HER2 scoring level, and an overall model-predicted HER2 score, consistent with ASCO/CAP guidelines. Model outputs are shown in **Figure 1A**.

### Model Evaluation

AIM-HER2 performance was assessed on an held-out test dataset consisting of additional slides obtained from five academic or commercial sources (N=804 total, 770 evaluable). Board-certified pathologists (N=52) with relevant experience provided manual HER2 scores based on ASCO-CAP guidelines, and AIM-HER2 scores were generated. Nested pairwise non-inferiority analysis^26^ was used to compare model performance to that of pathologists (N=3 pathologists per slide) via linearly-weighted Cohen’s kappa. In the nested pairwise framework, agreement among pairs of pathologists was compared in turn to agreement between AIM-HER2 and each pathologist, so that summary metrics account for inter-pathologist variability.

### HER2 Assisted Read Study

To evaluate the impact of AIM-HER2 on pathologist reads, we performed a cross-over study in which an additional independent set of WSIs (N=200) of HER2 IHC were scored by manual pathologist assessment and in an “AI Assist” fashion with AIM-HER2. These scores were compared to a consensus score derived from the manual scoring of an additional five pathologists (“reference consensus score”). As shown in **Figure 1B**, a study panel of 20 pathologists were divided into two cohorts, which determined the order of manual or AI-assisted scoring. After a three week washout period, the pathologists who originally scored manually then scored with AIM-HER2 assist, and the pathologists who originally scored using AIM-HER2 assist then scored manually. In order to minimize bias, WSI were presented to the pathologists in a random order during each scoring period.

As part of this study, the following endpoints were assessed: inter-rater agreement with and without AI, pathologist accuracy compared to consensus with and without AI, and model accuracy compared to consensus.

### Statistical Analysis

For the assisted read study, inter-pathologist agreement with and without assistance of AIM-HER2 was calculated via Krippendorff’s alpha for ordinal data . Accuracy among pathologists and AIM-HER2 against the consensus panel was quantified in terms of overall percent agreement (OPA), positive percent agreement (PPA), negative percent agreement (NPA), and Krippendorff’s alpha with ordinal weights. Correlations were quantified using Spearman’s rank correlation coefficient. The consensus score for each slide was defined as the median. All confidence intervals were generated by bootstrapping with 2000 iterations. All analyses were completed in the Python 3.10 programming language, using krippendorff, scipy, sklearn, pandas, and numpy packages.

## Results

### Development and validation of AIM-HER2

We hypothesized that a machine learning (ML) approach would have the potential to improve inter-pathologist agreement in providing HER2 scores. As such, we developed a machine-learning model (“AIM-HER2”) to generate accurate, slide-level HER2 scores aligned with ASCO-CAP guidelines in breast cancer HER2 IHC specimens with the goal of using this algorithm as an AI-assist tool.

AIM-HER2 performance was assessed on breast cancer slides (N=804 total, 770 evaluable) on which HER2 IHC was performed. Board-certified pathologists (N=52) with relevant experience provided manual HER2 scores based on ASCO-CAP guidelines. High concordance was observed between AIM-HER2-predicted and pathologist-labeled slide-level HER2 scores, both overall and for each scoring level (**Figure 2A-C**). Similar results were observed when assessing AIM-HER2 performance on multiple slide scanners (**Figure 2D**) and with multiple HER2 IHC antibody clones (**Figure 2E**).

**Figure 2.**
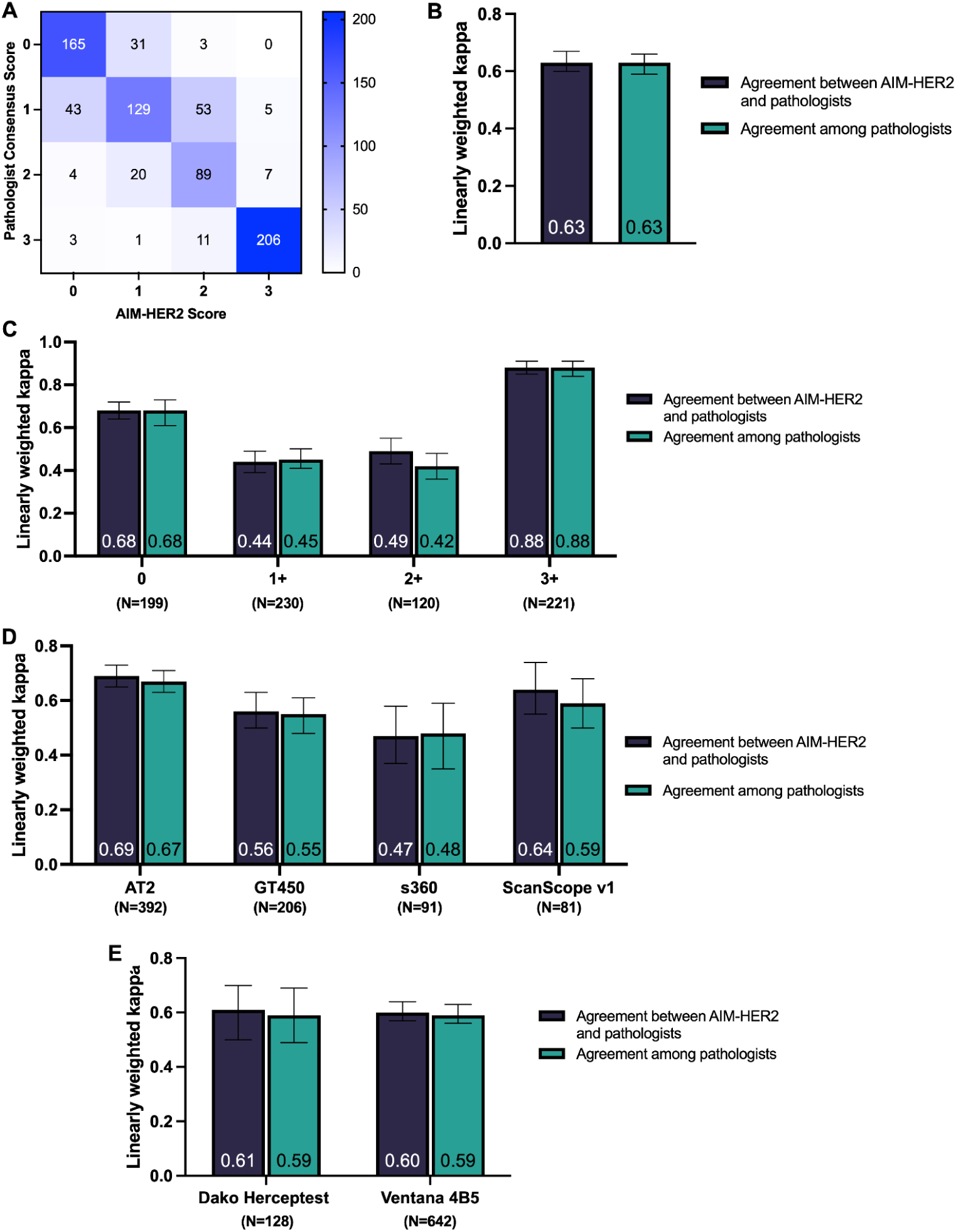
Concordance between AIM-HER2 predicted and pathologist-labeled slide-level HER2 scores for algorithm validation. A) Confusion matrix showing agreement between AIM-HER2 and pathologist consensus for each HER2 score. Linear kappa values were calculated to assess agreement between AIM-HER2 scores and pathologist scores as well as agreement among pathologists B) overall, C) for each HER2 score, D) for each slide scanner, or E) for each HER2 antibody clone using nested pairwise analysis.

### Inter-pathologist agreement for scoring of HER2 IHC

Having validated the performance of the AIM-HER2 algorithm, we sought to evaluate the impact of this model as an AI-assist tool for HER2 scoring in breast cancer. To do so, we conducted a retrospective reader study, in which HER2-trained pathologists (N=20) read a panel of cases (N=200) both with and without AI assistance, using a 2-cohort crossover design with a 3-week washout (**Figure 1B**). In addition, five expert HER2 pathologists read all 200 cases manually on a digital image platform to establish reference consensus scores.

First, we sought to understand the inter-pathologist agreement for manual HER2 scoring in our cohort. Previous work has revealed an opportunity to improve concordance when distinguishing between the scores of 0 and 1+ and between 1+ and 2+^10–13^. The agreement between the study pathologists and reference panel consensus in our cohort is shown in **Table 1**. Of the 200 cases assessed, 37.5% (N=75) had <70% of pathologists in agreement. Of these cases with low agreement, most had consensus scores of either 0 (N=26) or 1+ (N=36; total N=62/75, 82.67%). Across all cases, the average agreement between study pathologists was highly dependent on the consensus score, with the highest agreement observed in cases scored as 3+ (98%) and the lowest agreement observed in cases scored to be 1+ (66%) and 2+ (68%). The agreement in cases scored as 0 was observed to be 80%. These trends in agreement are consistent with those identified in previous studies^10–13^.

**Table 1.**
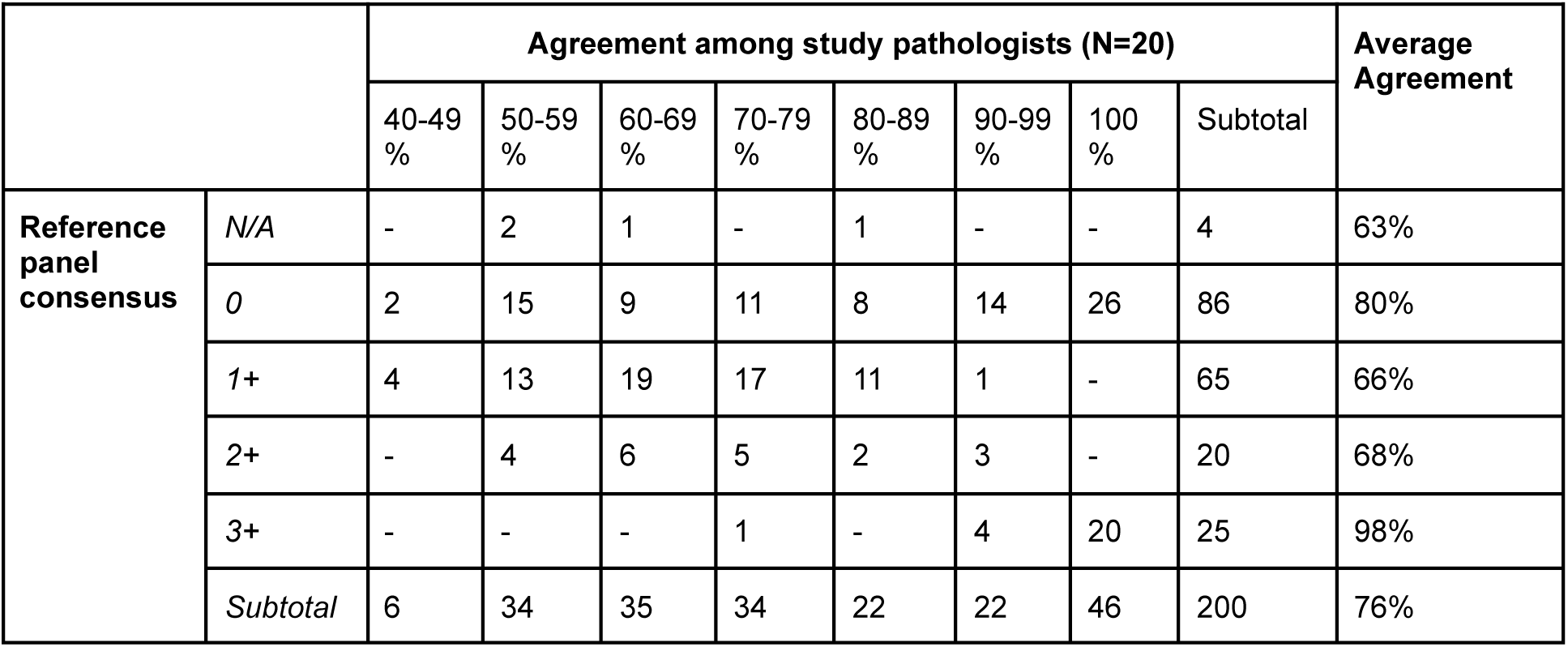
Percent agreement of study pathologists (N=20) compared to the consensus score of reference pathologists (N=5). The number of cases for each percent agreement range is shown.

To further understand the degree of inter-pathologist agreement in our cohort, we assessed the agreement among reference panel pathologists, as well (**Table 2**). Similar to the study panel, the agreement between reference panel pathologists was highly dependent on the consensus score, with the agreement being highest in cases scored as 3+ (98%) and lowest in cases scored as 1+ (81%) and 2+ (76%). Notably, the average agreement for the reference panel pathologists was equal to or higher than the average agreement for the study panel pathologists across all scores. However, for 25% of cases (N=50), only 2 or 3 (of 5) reference pathologists were in agreement. A comparison of the individual reference pathologist scores with the reference panel scores showed that the disagreement between reference pathologists tended to occur between the scores of 0 and 1+ and between 1+ and 2+ (**Table 3**).

**Table 2.** Percent agreement of reference pathologists (N=5) compared to the consensus score of the reference panel. The number of cases for each agreement range is shown.

|  |  | Agreement among reference panel pathologists |  |  |  |  | Average Agreement |
| --- | --- | --- | --- | --- | --- | --- | --- |
|  |  | 2 of 5 | 3 of 5 | 4 of 5 | 5 of 5 | Subtotal |  |
| Reference panel consensus | N/A | 4 | - | - | - | 4 | 40% |
|  | 0 | - | 18 | 13 | 55 | 86 | 89% |
|  | 1+ | - | 18 | 26 | 21 | 65 | 81% |
|  | 2+ | - | 9 | 6 | 5 | 20 | 76% |
|  | 3+ | - | 1 | 1 | 23 | 25 | 98% |
|  | Subtotal | 4 | 46 | 46 | 104 | 200 | 85% |

**Table 3.** Comparison of individual reference pathologist HER2 scores with consensus scores.

|  |  | Individual reference pathologist scores |  |  |  |  |
| --- | --- | --- | --- | --- | --- | --- |
|  |  | 0 | 1+ | 2+ | 3+ | Subtotal |
| Reference panel consensus | N/A | 35% (7) | 40% (8) | 20% (4) | 5% (1) | 100% (20) |
|  | 0 | 89% (381) | 10% (43) | <1% (3) | <1% (3) | 100% (430) |
|  | 1+ | 13% (42) | 81% (263) | 6% (20) | - | 100% (325) |
|  | 2+ | - | 17% (17) | 76% (76) | 7% (7) | 100% (100) |
|  | 3+ | - | - | 2% (3) | 98% (122) | 100% (125) |
|  | Subtotal | 43% (430) | 33% (331) | 11% (106) | 13% (133) | 1000 |

### Effect of AIM-HER2 on inter-pathologist agreement for scoring of HER2 IHC

To examine the effect of AIM-HER2 as an AI-assist tool, we compared the inter-pathologist agreements resulting from manual and AI-assisted HER2 scoring (**Figure 3**). AI-assistance with the algorithm improved inter-rater agreement by 6% across all HER2 scores (**Figure 3A**; p=0.001) and by 15% when comparing 0 vs 1+/2+/3+ (**Figure 3B**; p<0.0005). Inter-pathologist agreement of AIM-HER2 assisted reads was non-inferior (with a 0.1 margin) to manual assessment alone when comparing 0/1+ to 2+/3+ (**Figure 3C**; p<0.0005). Given the disagreement known to occur between the scores of 0 and 1+ and between 1+ and 2+^11^, we also examined the effect of AIM-HER2 assistance specifically at these levels. AI-assistance with AIM-HER2 significantly improved inter-rater agreement compared to manual assessment when assessing scores of 0 and 1+ (**Figure 3D**; p<0.0005) and scores of 1+ and 2+ (**Figure 3E**; p=0.006).

**Figure 3.**
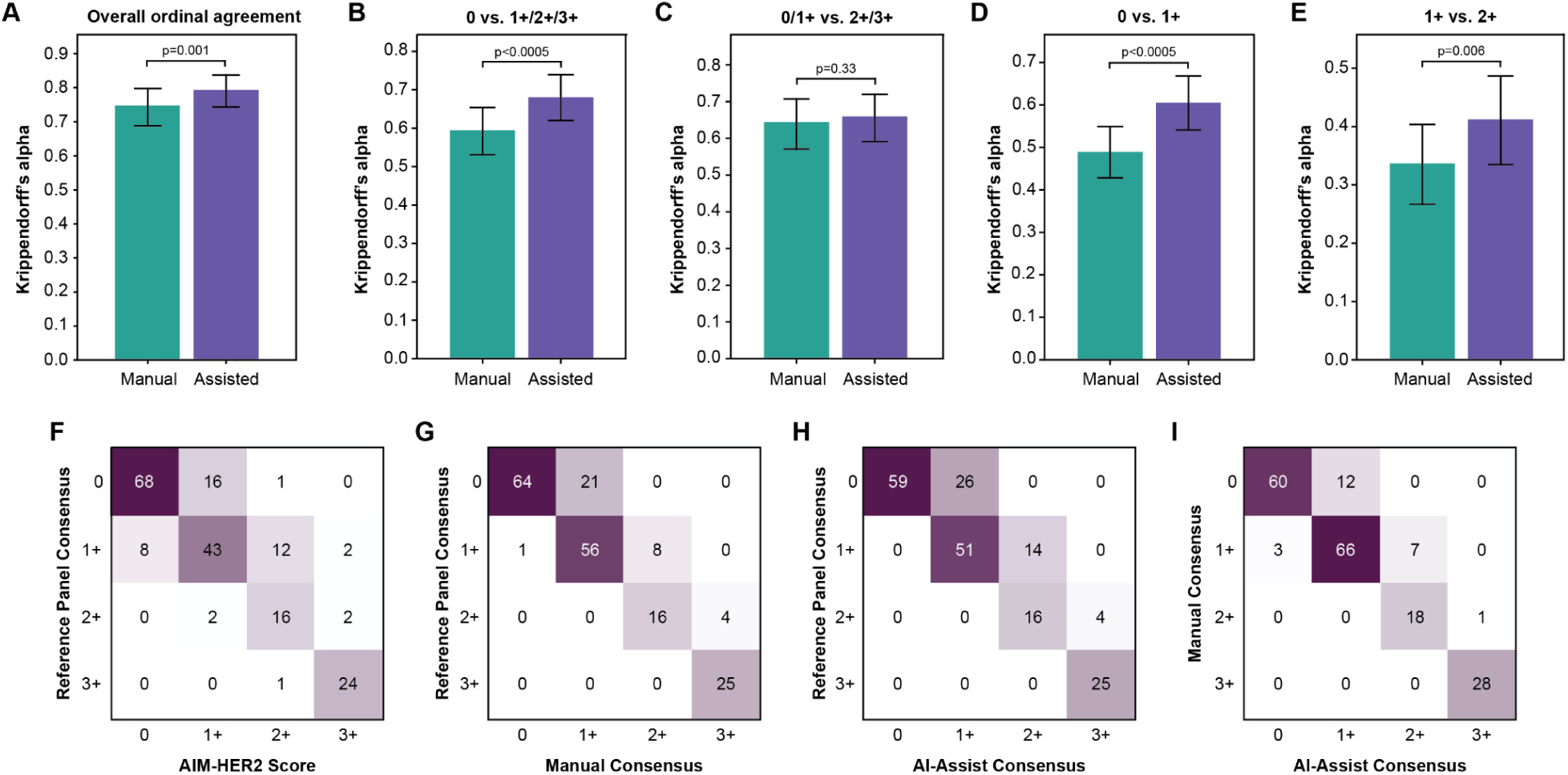
Comparison of manual and AIM-HER2-assisted HER2 scoring. Comparison of inter-pathologist agreements resulting from manual and AIM-HER2-assisted HER2 scoring A) across all scores and at the at cutoffs of B) 0 vs. 1+/2+/3+ and C) 0/1+ vs. 2+/3+.) Targeted analyses of inter-pathologist agreements were conducted at the D) 0 vs. 1+/2+/3+ and E) 0/1+ vs. 2+/3+ cutoffs. Comparisons of the reference consensus scores and F) AIM-HER2, G) manual consensus scores, and H) AIM-HER2-assisted consensus scores. I) Comparison of manual consensus scores with AI-assisted consensus scores.

Comparison of the HER2 scores predicted by AIM-HER2 and the reference panel consensus scores is shown in **Table 4** and **Figure 3F-H**. Interestingly, a comparison of the disagreement between AIM-HER2-predicted scores and the reference panel reveals a similar trend to the study panel of pathologists: both AIM-HER2 and study pathologists show an apparent bias toward overcalling relative to the reference panel. Interestingly, a higher concordance was observed between manual consensus and AI-assisted consensus (**Figure 3I**).

**Table 4.** Comparison of AIM-HER2-predicted HER2 scores with consensus scores.

|  |  | AIM-HER2 scores |  |  |  |  |
| --- | --- | --- | --- | --- | --- | --- |
|  |  | 0 | 1+ | 2+ | 3+ | Subtotal |
| Reference panel consensus | N/A | - | 1 | 1 | 2 | 4 |
|  | 0 | 80% (68) | 19% (16) | 1% (1) | - | 100% (85) |
|  | 1+ | 12% (8) | 66% (43) | 18% (12) | 3% (2) | 100% (65) |
|  | 2+ | - | 10% (2) | 80% (16) | 10% (2) | 100% (20) |
|  | 3+ | - | - | 4% (1) | 96% (24) | 100% (25) |
|  | Subtotal | 38% (76) | 31% (61) | 15% (30) | 14% (28) | 200 |

### Accuracy of AIM-HER2 as an AI-assist tool

When assessing the positive percent agreement (PPA) of AIM-HER2 assistance, we did not observe a significant difference across all HER2 scores (**Figure 4A**; p=0.83). However, AI-assistance led to significant improvements in PPA at the 0 vs. 1+/2+/3+ cutoffs (**Figure 4B**; p<0.0005) and at the 0/1+ vs. 2+/3+ cutoffs (**Figure 4C**; p<0.0005). In addition, decreased negative percent agreement (NPA) values with AI-assistance were also observed at the 0 vs. 1+/2+/3+ cutoffs (**Figure 4B**; p=0.005) and at the 0/1+ vs. 2+/3+ cutoffs (**Figure 4C**; p<0.0005). No significant difference in OPA was observed at the 0 vs. 1+/2+/3+ cutoff (**Figure 4B**; p=0.475), while a significant but small decrease in OPA was observed at the 0/1+ vs. 2+/3+ cutoff with AI-assistance (**Figure 4C**; p=0.017). When limiting our comparisons to 0 vs. 1+ and 1+ vs. 2+, no significant difference in OPA was observed between manual and AI-assisted scoring (**Figure 4D-E**). AI-assistance resulted in increased PPA at the 0 vs. 1+ cutoff (**Figure 4D**; p<0.0005) and the 1+ vs. 2+ cutoff (**Figure 4E**; p<0.0005). In addition, lower NPA was observed with AI-assistance at the 0 vs. 1+ cutoff (**Figure 4D**; p=0.007) and the 1+ vs. 2+ cutoff (**Figure 4E**; p=0.003).

**Figure 4.**
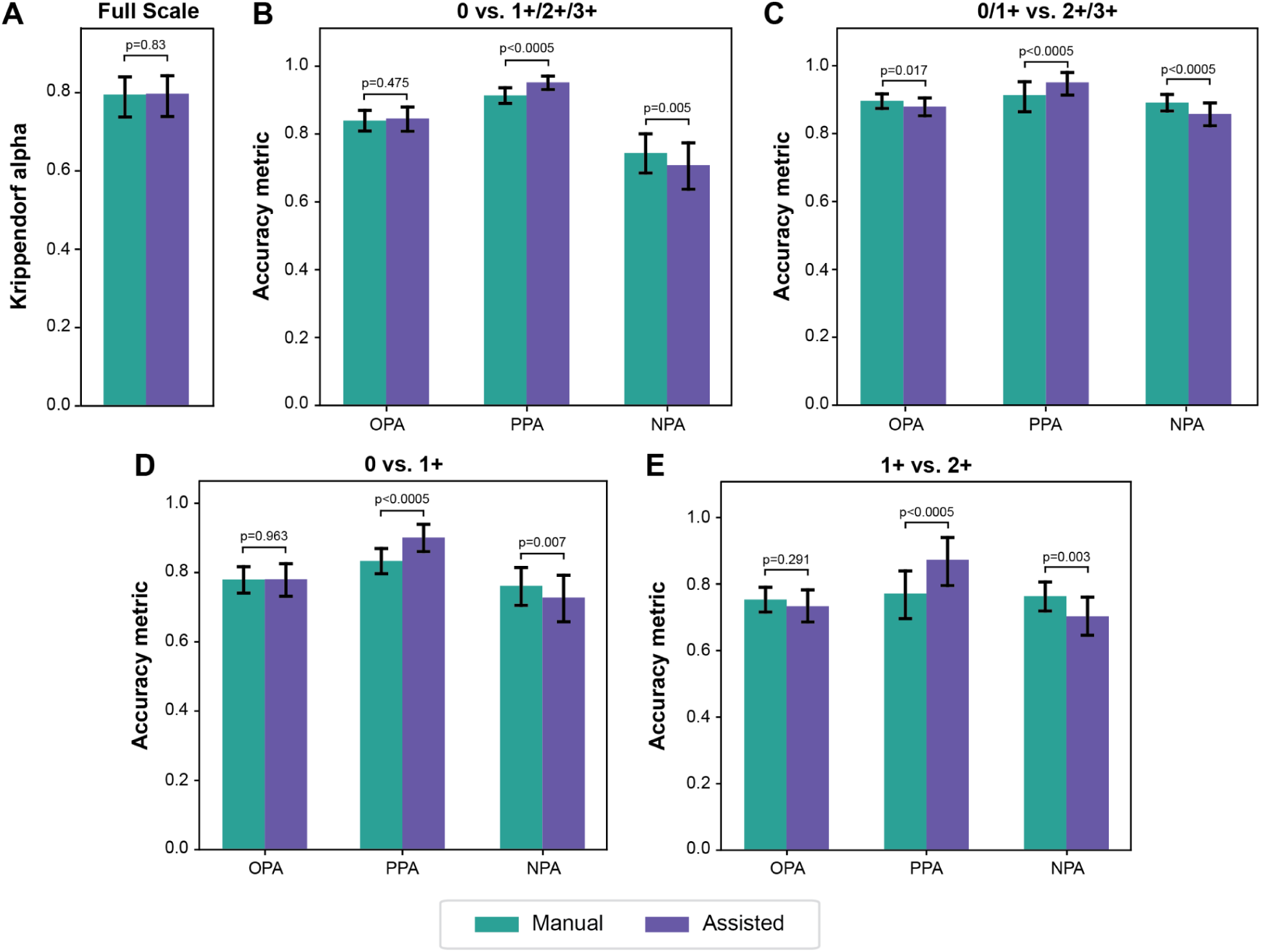
Metrics of AIM-HER2 accuracy. Metrics are shown A) across all HER2 scores as well as for the distinguishing scores at cutoffs of B) 0 vs. 1+/2+/3+, C) 0/1+ vs. 2+/3+, D) 0 vs 1+, and E) 1+ vs. 2+.

### Impact of AIM-HER2 at the individual case level

As summarized in **Table 5**, assessment of the relationship between the manual, AI, and AI-assisted scores reveals the “impact” of the algorithm on any individual case. When the manual read, AI read, and AI-assisted read are all concordant or all discordant with the reference score, the AI impact was considered neutral and AI-verified. The AI impact was considered positive if the manual score was discordant with the reference consensus while the AI and AI-assisted reads were concordant. In contrast, if the manual read is concordant with the reference consensus while the AI read and AI-assisted read were discordant, the AI impact was considered negative. An override of AIM-HER2 occurred when the AI-assisted HER2 score differed from the AIM-HER2 score. In such instances, a correct override occurred when the AI-assisted score was concordant with the reference consensus score, and an incorrect override occurred when the AI-assisted score was discordant with the reference consensus score.

**Table 5.** Impact of AIM-HER2 by reference consensus score.

| Manual Score vs. Reference Consensus | AIM-HER2 Score vs. Reference Consensus | AI-Assisted Score vs. Reference Consensus | AI Impact | Reference Score |  |  |  |  |
| --- | --- | --- | --- | --- | --- | --- | --- | --- |
|  |  |  |  | 0 (N=85) | 1+ (N=65) | 2+ (N=20) | 3+ (N=25) | Subtotal Reads (N=3900) |
| Concordant | Concordant | Concordant | Neutral (AI-Verified) | 1027 (60%) | 453 (35%) | 192 (48%) | 473 (95%) | 2145 (55%) |
|  |  | Discordant | Neutral (Incorrect override) | 102 (6%) | 110 (8%) | 24 (6%) | 3 (1%) | 239 (6%) |
|  | Discordant | Concordant | Neutral (correct override) | 52 (3%) | 151 (12%) | 18 (4%) | 13 (3%) | 234 (6%) |
|  |  | Discordant | Negative | 84 (5%) | 111 (9%) | 13 (3%) | 2 (0%) | 210 (5%) |
| Discordant | Concordant | Concordant | Positive | 103 (6%) | 171 (13%) | 62 (16%) | 4 (1%) | 340 (9%) |
|  |  | Discordant | Neutral (Incorrect override) | 128 (8%) | 126 (10%) | 42 (10%) | 0 (0%) | 296 (8%) |
|  | Discordant | Concordant | Neutral (correct override) | 22 (1%) | 49 (4%) | 7 (2%) | 1 (0%) | 79 (2%) |
|  |  | Discordant | Neutral (AI-Verified) | 192 (11%) | 129 (10%) | 42 (10%) | 4 (1%) | 357 (9%) |

We assessed the degree to which AI impacted the HER2 score, as well as the degree to which an override occurred. These results are summarized **Table 5** and **Figure 5A**. Across all AI-assisted reads (N=3,900), a neutral impact was observed in 64% of cases (N=2,502). In total, AI impacted 14% of reads (N=550), with a positive impact observed in 8.7% of total reads (N=340) and a negative impact observed in 5.4% of total reads (N=210). Reads with negative impact occurred most frequently at scores of 0 and 1+, while reads with positive impact occurred most frequently at scores of 0, 1+, and 2+ (**Table 5**).

**Figure 5.**
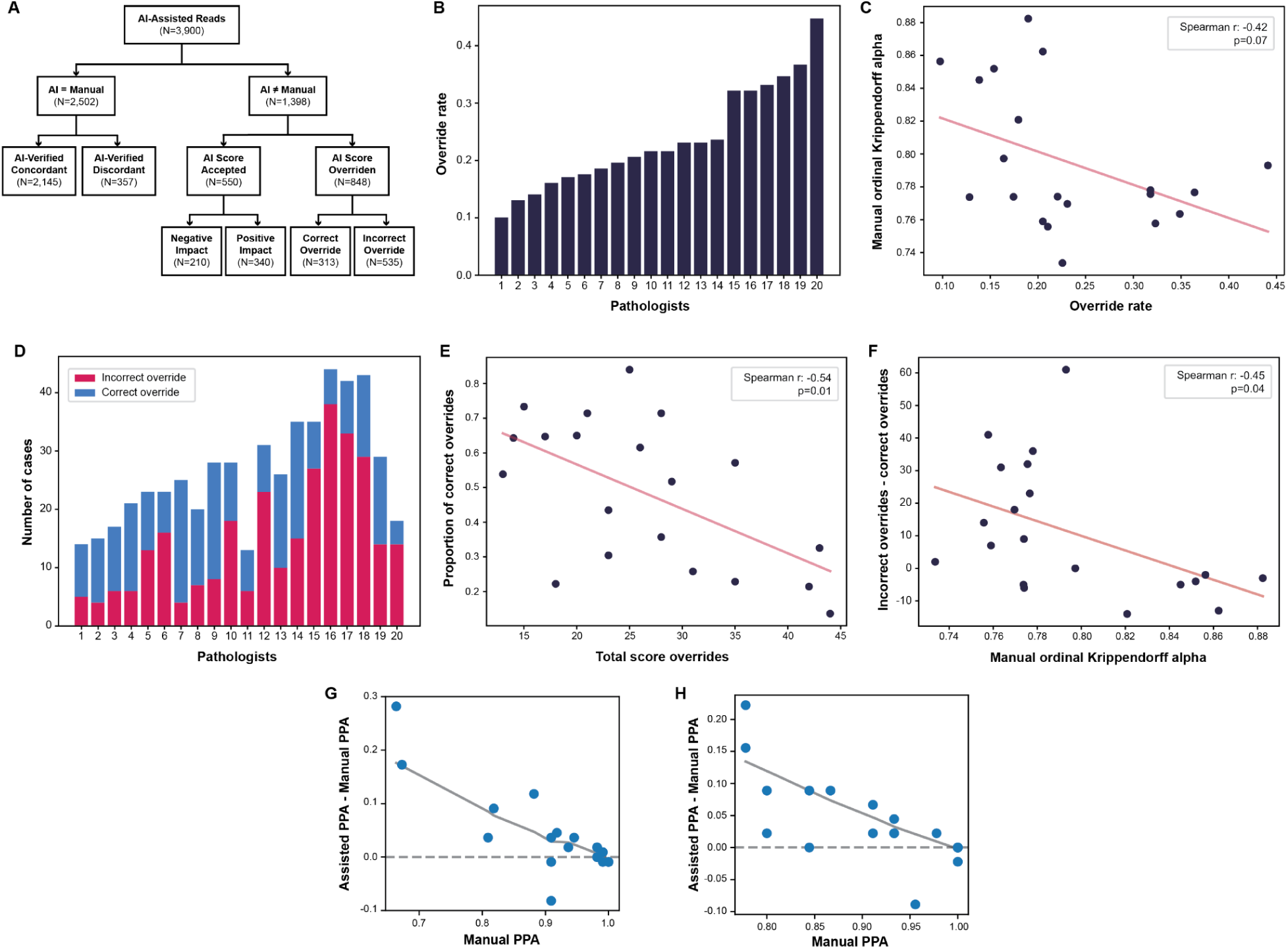
Pathologist override of AIM-HER2 scores. A) Proportion of total reads accepted, overridden, or agreeing with the manual score. B) Rate at which each study pathologist overrode the AIM-HER2 score. C) Association between the rate at which study pathologists overrode the AIM-HER2 score and the pathologists’ manual accuracy compared to the reference panel. D) Proportion of correct overrides for each study pathologist. E) Association between the proportion of correct overrides with the total number of overridden scores. F) Association between pathologist manual accuracy and proportion of correct overrides. G-H) Association between improvement in PPA with manual PPA for the G) 0 vs. 1+/2+/3+ cutoff and the H) 0/1+ vs. 2+/3+ cutoff

Among individual study pathologists, observed override rates varied widely, from 10% to 45% with a median of 21.6% (**Figure 5B**). To better understand this level of variance, we evaluated the association of study pathologists’ manual accuracy to their override rates. Pathologist override rates were observed to negatively associate with agreement to the manual reference panel (**Figure 5C**; Spearman r = -0.42; p = 0.07). The ratio of incorrect overrides to correct overrides also varied widely among study pathologists, with some pathologists showing a high proportion of incorrect overrides (**Figure 5D**). The rate of incorrect pathologist overrides was negatively associated with both the total number of overridden scores (**Figure 5E**; Spearman r = -0.54; p=0.01) and agreement to the manual reference panel (**Figure 5F**; Spearman r = -0.45; p=0.04).

The association between a pathologist’s manual accuracy and benefit from AI assistance was also examined. For the 0 vs. 1+/2+/3+ cutoff (**Figure 5G**) and 0/1+ vs. 2+/3+ cutoff (**Figure 5H**), the greatest benefit to PPA (assisted PPA - manual PPA) was observed for the pathologists with the lowest manual PPA, suggesting that pathologists with lower manual accuracy benefit most from AI assistance. Minimal effects of AI assistance on OPA and NPA were observed for the 0 vs. 1+/2+/3+ cutoff (**Supplemental Figure 2A,B**) and 0/1+ vs. 2+/3+ cutoff (**Supplemental Figure 2C,D**).

### Dependency of reference panel on scoring accuracy

While the overall inter-reader agreement for our reference panel was observed to be high (85%), we did note a dependence of the agreement on the consensus score. In addition, we observed a relatively high number of cases where there was high disagreement among the panel (**Tables 2, 3**). Therefore, we sought to further assess the impact of the reference panel performance on accuracy metrics for manual and AI-assisted HER2 scoring.

To address this question, we collected AI-assisted HER2 scores from our reference panel such that we had manual and AI-assisted HER2 scores from 25 pathologists (study pathologists and reference pathologists). Using all 25 pathologists as a reference panel, the concordance of AIM-HER2 with the larger reference panel (**Figure 6A**) resembled that from our original five-pathologist reference panel (**Figure 3F**).

**Figure 6.**
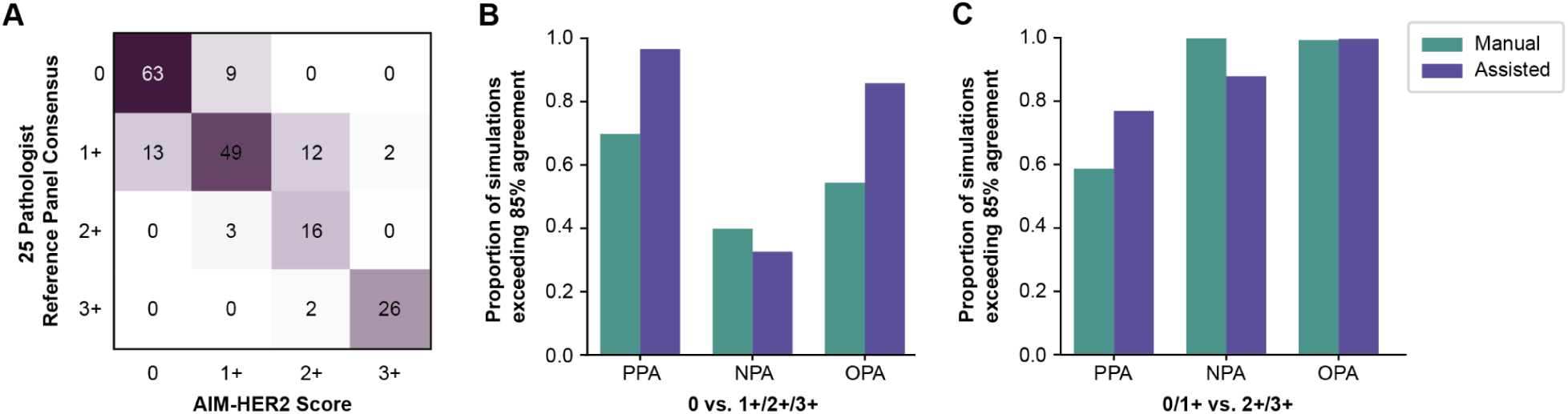
Effect of reference panel composition on accuracy metrics for HER2 scoring. A) Comparison of AIM-HER2 scores with consensus scores from a 25-pathologist reference panel. B-C) A simulation was performed to generate 500 unique reference panels, and the accuracy of manual and AI-assisted was calculated for each iteration. The proportion of simulations for which positive predictive, negative predictive, and overall predictive accuracy metrics were above 85% are shown for B) 0 vs. 1+/2+/3+ and C) 0/1+ vs. 2+/3+ cutoffs.

In addition, from the reads provided by all 25 pathologists, a simulation was performed in which a series of distinct 5-pathologist reference panels (N=500) were constructed. For each of these 500 reference panels, accuracy metrics were computed. A comparison of the accuracy metrics calculated by comparing to each reference panel revealed large ranges in OPA, PPA, and NPA for AIM-HER2 accuracy (**Supplemental Figure 3A)**, manual pathologist accuracy (**Supplemental Figure 3B**), and AI-assisted pathologist accuracy (**Supplemental Figure 3C**). At the 0/1+ cutoff, the median of the average manual OPA, PPA, and NPA values were 0.89 (range: 0.75 to 0.92), 0.92 (range: 0.60 to 0.99), and 0.85 (range: 0.62 to 1). At the 1+/2+ cutoff, the median of average manual OPA, PPA, and NPA values were 0.93 (range: 0.78 to 0.97), 0.94 (range: 0.74 to 1), and 0.95 (range: 0.72 to 1).

Given the observed variability in accuracy with different reference panels, we sought to understand the impact of AI-assistance on HER2 scoring accuracy independent of the reference panel. To do so, we used the reference panel simulations to determine the proportion of accuracy metrics that would be greater than 85% in value. For comparisons of HER2 scores of 0 versus 1+/2+/3+, a greater proportion of analyses exceeded 85% PPA and OPA with AI assistance than with manual scoring, while the number of NPA simulations exceeding 85% was slightly higher with manual scoring (**Figure 6B**). For comparisons of HER2 scores of 0/1+ versus 2+/3+, a greater proportion of analyses exceeded 85% for PPA with AI assistance, the number of NPA simulations exceeding 85% was higher with manual scoring, and the proportion of analyses were equivalent for OPA (**Figure 6C**).

## Discussion

Given that HER2 score is the key determinant of patient eligibility for HER2-targeted therapies such as trastuzumab and trastuzumab deruxtecan, accurate scoring is of the utmost clinical importance. Computer vision approaches have been proposed to mitigate the issue of reproducibility in diagnostic pathology. Here, we describe AIM-HER2, an algorithm designed to predict ASCO/CAP HER2 immunohistochemistry scores in breast cancer specimens.

As a standalone model for HER2 score prediction, pairwise AIM-HER2-pathologist and pathologist-pathologist agreement rates were similar. This result is upheld when slides were imaged using multiple scanning platforms and stained using multiple HER2 antibody clones, suggesting that AIM-HER2 would generalize well across these clinical variables. Given the performance and generalizability of AIM-HER2, we also examined the algorithm as an AI-assist tool, where pathologists use an AI algorithm to guide clinical decision making ^24^. Given the recent finding that patients with HER2-low disease may be a clinically relevant population ^9, 27, 28^, accurate HER2 scoring is necessary to ascertain that patients receive the most appropriate treatment. However, high levels of intra- and inter-pathologist variability for HER2 manual scoring, particularly at the 0/1+ and 1+/2+ cutoffs, have been observed, potentially leading to incorrect scores in a subset of patients ^10–16^. These results emphasize the need for improved accuracy and consistency in HER2 scoring, particularly around the 0/1+ and 1+/2+ scoring levels. In the AI-assist setting, AIM-HER2 significantly increased inter-pathologist agreement across all scores, but especially at the 0/1+ and 1+/2+ cutoffs. Importantly, AI-assist using AIM-HER2 significantly increased PPA at these cutoffs, as well. These results support the continued investigation of AIM-HER2 as an AI-assist tool, especially for the scoring of cases for which manual scoring may yield equivocal or incorrect results.

While AI assistance has been shown to improve the reproducibility in multiple common grading and scoring tasks ^29–32^, trends regarding how pathologists interact with AI-assist tools, including the degree to which they are in agreement with such algorithms, have yet to be explored. Here, we assessed the interaction between pathologists and AIM-HER2 when used for AI assistance. Perhaps not surprisingly, a wide range of overall override rates were observed across the study pathologists, as well as high variability in each pathologist’s override accuracy. Of the 3900 total AI-assisted reads amassed in this study, disagreements with the AIM-HER2 score occurred for roughly 22% of reads. At the individual pathologist level, override rates ranged from 10-45% (i.e., acceptance rates ranged from 55-90%). These override rates, as well as the frequency of incorrect overrides and benefit from AI assistance, were correlated with manual pathologist accuracy. Overall, 63% of overrides were incorrect (e.g., a pathologist changing a correct AIM-HER2 score to an incorrect score). While we observed that AI assistance most benefitted pathologists with lower manual PPA, a similar study that includes a final manual read from all pathologists would be needed to fully confirm this finding as a control. These results, combined with the overall improvements in HER2 scoring reproducibility, suggest that AI assist tools can be a valuable asset to pathologists, but specific training for using an AI assist workflow, as well as improved trust in AI tools, may be necessary. Improved trust in AI assistance in medicine may also be required for these tools to reach their full potential. A recent study assessing the benefit of a large language model (GPT-4) assistance for diagnostic reasoning revealed a propensity of physicians to ignore the model’s guidance ^33^, while radiologists were more likely to accept the recommendation of a fellow radiologist rather than an AI-derived source^34^.

In addition, examination of the results from our reference panel revealed that even within that group, a high degree of discrepancy was observed. For 25% of cases (N=50), only 2 or 3 (of 5) reference pathologists were in agreement. A comparison of the individual reference pathologist scores with the reference panel scores showed that the disagreement between reference pathologists tended to occur between the scores of 0 and 1+ and between 1+ and 2+, in line with previous literature around these scoring cutoffs ^10–16, 35^. The root causes of these observed scoring discrepancies remain unclear and should be addressed in future studies, perhaps with a focus on differences in visual search patterns between pathologists, as has been done in other domains ^36, 37^. Still, the lack of true consensus between the reference pathologists at these scoring levels has clear clinical implications: if the reference panel had consisted of three pathologists instead of five, as many as 25% of cases (N=50) could have been assigned a different score, potentially changing the treatment recommendations for the patients in question. As such, we performed a simulation to generate 5-pathologist reference consensus scores from 500 combinations of the pathologists included in our study, and we compared AIM-HER2 metrics to each. Notably, we observed a wide range of OPA, PPA, and NPA scores in this analysis. These results strongly suggest that, when comparing a scoring algorithm to a reference consensus score, the composition of the reference panel has significant implications for the resulting accuracy metrics. Recent results also suggest that, the optimal number of reference pathologists to maximize reproducibility depends on the individual IHC biomarker: while tests measuring consistently expressed proteins such as estrogen receptor (ER) and progesterone receptor (PR) may only necessitate 2-3 reference pathologists, tests measuring more variable markers, such as Ki67, may necessitate up to five reference pathologists ^38^.

Further work is needed to identify the optimal reference panel composition for adjudicating HER2 scoring models. Given potential biases in reference panels, such as that observed here, a complementary approach – such as nested pairwise comparison – should be examined to avoid the improper assumption that consensus values are infallible. Perhaps, for example, ranking how an AI tool performs relative to pathologists as a percentile would be a more practical lens through which to evaluate AI accuracy. Additional study is also necessary to better understand how these results may be impacted by the specific user interface, as well as how pathologists choose to interact with the AI.

In conclusion, this study demonstrates that the use of AIM-HER2 as an AI-assist tool for scoring HER2 IHC in breast cancer may improve pathologist reproducibility and accuracy, particularly at the 0/1+ and 1+/2+ cutoffs. This AI-assist tool has the potential to aid in the accurate determination of HER2 scores, enabling future translational and clinical research in oncology.

## Data Availability

Access to HER2 prediction heatmaps is available upon reasonable request to academic investigators without relevant conflicts of interest for non-commercial use who agree not to distribute the data. Access requests can be made to.

## Supplemental Figures

**Supplemental Figure 1.**
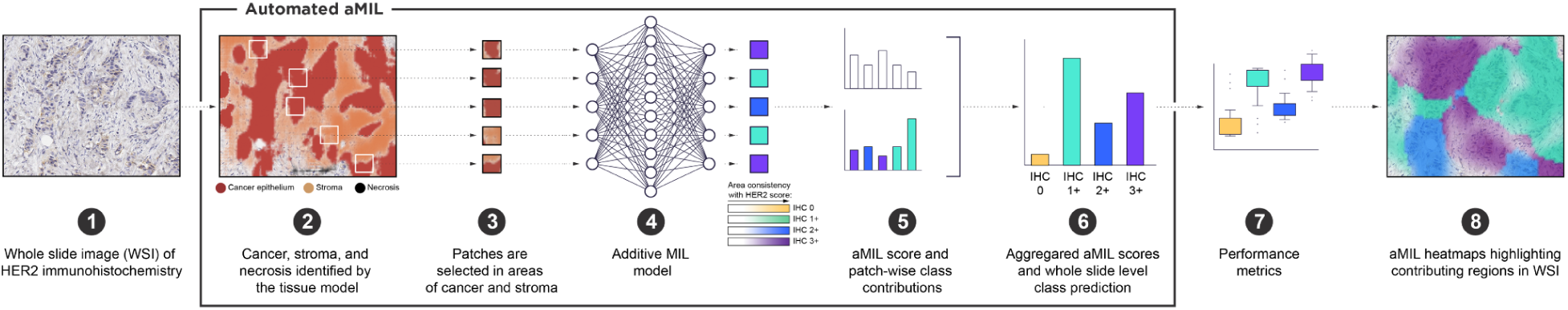
AIM-HER2 aMIL architecture. From WSI of HER2 IHC, image patches are selected in model-predicted areas of cancer and stroma. The aMIL model predicts the likelihood of each patch as being a certain HER2 score (IHC 0, 1+, 2+, or 3+, consistent with ASCO/CAP guidelines). From the model output, a slide-level HER2 score can be predicted, and image overlays show the relative contribution of areas of tissue to the model-predicted score.

**Supplemental Figure 2.**
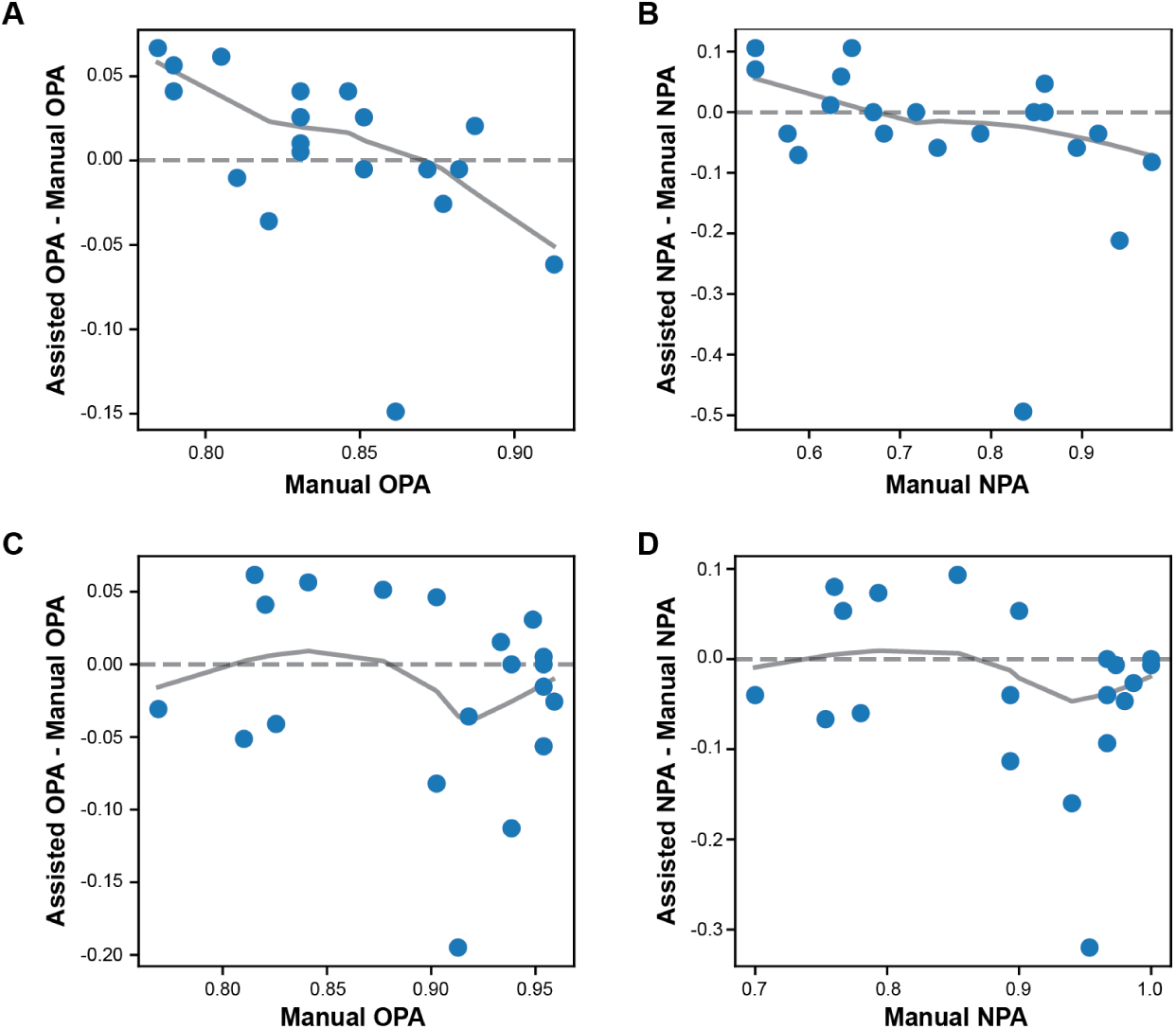
Association between pathologist manual accuracy and accuracy metrics.

**Supplemental Figure 3.**
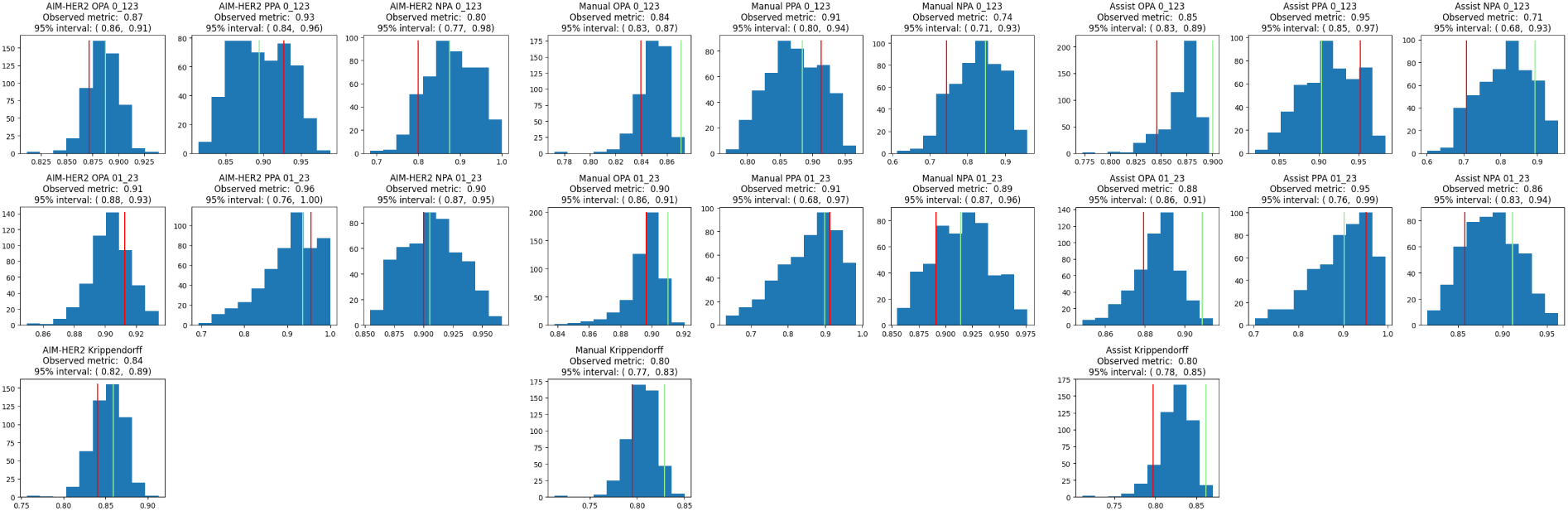
Accuracy metrics after reference panel simulation. OPA, PPA, and NPA for A) AIM-HER2, B) manual scoring, and C) AI-assisted scoring are shown after a simulation of reads using 500 distinct reference consensus panels. The red bar indicates the metrics for the original 5-pathologist reference panel, and the green bar indicates the average of per-pathologist metrics when the reference panel consists of all 25 pathologists.

